# Safety and Immunogenicity of Omicron Protein Vaccines in mRNA-Vaccinated Adolescents: A Phase 3, Randomised Trial

**DOI:** 10.1101/2024.11.07.24316926

**Authors:** Chijioke Bennett, Gordon Chau, Erika Clayton, Laurence Chu, Jacqueline Alvarez, Ausberto B Hidalgo, Khozema Palanpurwala, Joyce S. Plested, Mingzhu Zhu, Shane Cloney-Clark, Zhaohui Cai, Raj Kalkeri, Karim Hegazy, Katherine Smith, Susan Neal, Fernando Noriega, Raburn M. Mallory, Jeffrey M. Adelglass 2019nCoV-314 Study Investigators

**Affiliations:** Novavax, Inc., 700 Quince Orchard Road, Gaithersburg, Maryland, 20878, USA; DM Clinical Research, 7411 Lake Street #L120, River Forest, Illinois, 60305, USA; Benchmark Research, 3100 Red River Street #1, Austin, Texas, 78705, USA; ITB Research, 9220 Sunset Drive, Suite 105, Miami, Florida, 33173, USA; Alfa Medical Research, 7777 Davie Road Extension #202a, Davie, Florida, 33024, USA; DM Clinical Research, 7908 North Sam Houston Parkway W #200, Houston, Texas, 77064, USA; Research Your Health, 6020 West Parker Road, Suite 430, Plano, Texas, 75093, USA

**Keywords:** COVID-19, SARS-CoV-2, booster, variant

## Abstract

**Objectives:** Safety and immunogenicity assessment of updated monovalent and bivalent SARS-CoV-2 vaccines in adolescents.

**Methods:** This phase 3, double-blinded study randomised 12–<18-year-old participants, who received ≥2 prior doses of an approved/authorised mRNA-based COVID-19 vaccine, 1:1 to receive NVX-CoV2601 (XBB.1.5) or a bivalent vaccine (NVX-CoV2373 [Wuhan] + NVX-CoV2601). The primary immunogenicity endpoint was day-28 neutralizing antibody (nAb) geometric mean titres (GMTs) against XBB.1.5. Safety endpoints were solicited reactogenicity ≤7 days and unsolicited adverse events (AEs) ≤28 days post vaccination and frequency/severity of predefined AEs of special interest through day 180.

**Results:** Of 401 randomised participants, nAb GMTs against XBB.1.5 increased (GMFR [95% CI]) for both NVX-CoV2601 (12.2 [9.5–15.5]) and the bivalent vaccine (8.4 [6.8–10.3]); post-vaccination responses to ancestral SARS-CoV-2 and the JN.1 variant were also observed. Increases in anti-spike IgG levels were comparable between the groups. Solicited and unsolicited AEs were mild to moderate, with similar occurrence among the groups. Severe and serious events were rare and unrelated to the study vaccines; no PIMMCs or myocarditis/pericarditis were reported.

**Conclusions:** NVX-CoV2601 elicited more robust antibody responses to XBB.1.5 and ancestral virus, compared with a bivalent formulation. The safety profile within each group was consistent with the prototype vaccine (NVX-CoV2373).

## Introduction

Prototype vaccines (ie, mRNA-1273, BNT162b2, and NVX-CoV2373) to ancestral severe acute respiratory syndrome coronavirus 2 (SARS-CoV-2) played a significant role in the early management of the COVID-19 pandemic.^1–3^ Rapid evolution and widespread circulation of viral variants has resulted in immune evasion and decreased immunity in recipients of prototype vaccines.^4–6^ In May and June of 2023, regulatory agencies (ie, the World Health Organisation [WHO], the European Medicines Agency [EMA], and the Vaccines and Related Biological Products Advisory Committee of the United States [US] Food and Drug Administration [FDA]) had regional meetings to discuss updates to COVID-19 vaccines and provided direction that formulations be monovalent compositions directed to XBB.1.5.^7–9^ This guidance was primarily based on nonclinical data for cross-neutralising antibody production from XBB.1.5-based vaccines across XBB subvariants^8,10,11^ and was partially based on evidence that variant strain mutations in the XBB sublineages were preserved.^10^ Additional support for XBB as a reasonable vaccine target is that this lineage of variants has demonstrated the ability to evade neutralising antibodies from both natural immunity and those induced by prior versions of COVID-19 vaccines.^12–14^

NVX-CoV2601 is an adjuvanted nanoparticle vaccine that contains recombinant spike (rS) protein from XBB.1.5 and Matrix-M™ adjuvant. Matrix-M is a saponin-based adjuvant shown to stimulate immune responses across a variety of vaccines.^15,16^ NVX-CoV2601 was generated based on similar protein technology used for the authorised vaccine (NVX-CoV2373) that contains the rS of ancestral SARS-CoV-2. Using clinical data from the prototype vaccine and preclinical data for the new formulation, NVX-CoV2601 was authorised for use in those aged ≥12 years by the EMA, the US FDA, and the WHO in the fall of 2023,^17–19^ and is supported by clinical findings in adults.^20^

Based on the rapid evolution of SARS-CoV-2 subvariants and shifts in recommendations from authorities, future vaccine recommendations have the potential to be monovalent (as per current guidance)^7–9^ or bivalent (as with the ancestral and BA.5 combination in 2022). Clinical immunogenicity and safety data in adolescents for variant-based vaccines, particularly XBB.1.5, are limited.^21,22^ As vaccines are updated, it is critical to continue gathering immunogenicity and safety data, across populations, not only for regulatory compliance, but also to inform the development of future vaccines. The 2019nCoV-314/NCT05973006 phase 3, randomised, double-blind trial was conducted to evaluate the safety and immunogenicity of the XBB.1.5-directed vaccine, NVX-CoV2601, in its monovalent form and as a bivalent combination with the prototype vaccine (NVX-CoV2601 + NVX-CoV2373). To provide additional data in adolescents, who are part of the indicated population for the vaccine, a targeted population of 12- to <18-year–olds in the US who had received ≥2 doses of an mRNA-based COVID-19 vaccine (mRNA-1273 or BNT162b2) were enrolled.

## Methods

### Study design and participants

The phase 3, randomised, observer-blinded 2019nCoV-314/NCT05973006 study enrolled previously vaccinated adolescents to evaluate the safety and immunogenicity of a single dose of nanoparticle monovalent (NVX-CoV2601 [XBB.1.5 rS]) and bivalent (NVX-CoV2601 plus NVX-CoV2373 [ancestral SARS-CoV-2 rS]) vaccines containing SARS-CoV-2 rS protein and adjuvanted with Matrix-M™ (Figure S1). Participants were medically stable 12- to <18-year-olds screened across 20 sites in the US and had received ≥2 prior doses of an approved/authorised mRNA vaccine ≥90 days prior to study vaccination. Key exclusion criteria were receipt of other investigational vaccines ≤90 days or an influenza vaccine ≤14 days before study vaccination, ongoing immunomodulatory therapy, chronic administration of immune-modifying drugs ≤90 days of study vaccination, or a history of myocarditis/pericarditis. Other vaccines recommended for 12- to <18-year–olds were allowed, as medically indicated. Participants found to be SARS-CoV-2 positive during the study were not excluded from participation. Informed consent was collected for each participant.

Participants were randomised 1:1 on day 0 per an interactive web response system and stratified by number of prior COVID-19 vaccinations. Study personnel were blinded to vaccine assignment, other than predetermined individuals who managed vaccine logistics (eg, preparation, administration), and did not have a role in study-related assessments or data collection. Participants were unblinded after the end of the study. The trial protocol can be found in the Supplementary material.

### Procedures

Participants were screened up to 14 days before study vaccine administration (day 0); if feasible, screening and day 0 could be the same day (Figure S1). On day 0, prior to study vaccine administration, nasal swabs were collected to perform qualitative polymerase chain reaction (PCR) for SARS-CoV-2 infection and blood samples for immunogenicity testing. Immunogenicity was assessed through these validated assays: anti-rS IgG^23^ enzyme-linked immunosorbent assay (ELISA) and pseudovirus-based neutralising antibody assay for XBB.1.5 and the ancestral SARS-CoV-2 strain.^24^ Participants received a single intramuscular injection of NVX-CoV2601 (5 µg rS) or a bivalent vaccine (NVX-CoV2601 + NVX-CoV2373; 2.5 µg rS of each), which were formulated with 50 µg of Matrix-M™ adjuvant. There was a minimum 15-min observation period to monitor for any immediate hypersensitivity/anaphylaxis reactions. Follow-up visits occurred in person on days 28, 90, and 180 (end of study) and via phone on days 56, 118, and 146. Safety was assessed throughout the study. Additional blood samples for immunogenicity tests were collected during in-person visits.

### Outcomes

Participants were trained to use an eDiary to record any solicited reactogenicity events post study vaccination, from day 0 through day 6. Solicited local (ie, pain, tenderness, redness, or swelling) and systemic (ie, fever, nausea/vomiting, headache, fatigue/malaise, muscle pain, and joint pain) treatment-emergent adverse events (TEAEs) were recorded in the eDiary. Unsolicited TEAEs (incidence, severity, and relation to vaccine) were collected through 28 days post vaccination. These included adverse events of special interest (AESI), serious adverse events (SAEs), and medically attended adverse events (MAAEs). AESIs, SAEs, and related MAAEs were collected through day 180. TEAEs were coded according to system organ class and preferred term per the Medical Dictionary for Regulatory Activities version 26.0. Investigators documented TEAE severity and assessed relation to study vaccine. AESI included potentially immune-mediated medical conditions (PIMMCs), myocarditis/pericarditis, and complications specific to COVID-19. Immunogenicity was investigated in each vaccine group by assessing neutralising antibody and anti-rS IgG responses to XBB.1.5 and the ancestral strain through validated assays.^23,24^ Exploratory outcomes included immunogenicity responses to the JN.1 variant.

### Statistical analyses

Sample size was based on clinical and practical considerations and not on a formal statistical power calculation. With 200 participants in each treatment group, the probability to observe at least one participant with a TEAE was >99.9%, if the true incidence of the TEAE was 5% (86.6% probability if the true incidence was 1%). This study was not powered for cross-group comparisons of noninferiority or superiority.

The primary safety objective was assessed through post-study vaccination endpoints of reactogenicity through day 6; unsolicited TEAEs through day 28; and AESI, SAEs, and related MAAEs through day 180. The safety analysis sets included all participants who provided consent, were randomised, received at least one dose of study vaccine, and were analysed per treatment actually received. All safety analyses were descriptive, with the number and percentage of participants recorded based on highest degree of TEAE severity and relatedness to the study vaccine; 95% CIs were calculated using the Clopper–Pearson method.

The primary immunogenicity objective was to describe the neutralising antibody response of each vaccine group against XBB.1.5 via primary endpoints of geometric mean titres (GMTs, ID_50_) at day 28 and geometric mean fold rise (GMFR) in GMTs from day 0 to day 28. Secondary immunogenicity endpoints included assessment of the neutralising antibody response and seroresponse rate (SRR) at days 90 and 180 as well as anti-rS IgG geometric mean ELISA units (GMEUs) and associated outcomes at days 0, 28, 90, and 180. Assessments included GMFRs and SRRs at days 28, 90, and 180, compared with day 0. The per-protocol analysis sets included all participants who received the prescribed study vaccine; had serology results for both day 0 (baseline) and another time point being analysed; were PCR negative at baseline for SARS-CoV-2; and had no major protocol violations or events that might have impacted the immunogenicity response (see Supplementary material for more details on protocol deviations). A per-protocol analysis set was determined for each immunogenicity assay and study visit. This study was not designed (and samples size was not powered) for formal statistical evaluation of immunogenicity.

GMTs/GMEUs and corresponding 95% CIs were summarised by vaccine group. GMTs were calculated as the antilog of the mean of log-transformed titre values and GMFR as the antilog of the mean of log-transformed fold-rises. The 95% CIs were calculated based on the *t*-distribution of the log-transformed GMT or GMFR, then back transformed to the original scale. Between-group GMT ratios (GMTRs) and the two-sided 95% CIs were computed using the analysis of covariance with the vaccine group as the fixed effect and the titre at day 0 (ie, adjusted for intergroup variation in baseline [pre-vaccination] titres) as the covariate. The mean difference between vaccine groups and the corresponding CI limits were exponentiated to obtain the GMTRs and the corresponding 95% CIs. Seroresponse was defined as a ≥4-fold increase in post-vaccination titre from baseline (or from the lower limit of quantification if the baseline value was below this limit). SRR and SRR difference 95% CIs were calculated based on the Clopper–Pearson exact and Miettinen–Nurminen methods, respectively. Anti-rS IgG GMEUs, GMFRs (compared with day 0), and 95% CIs were summarised.

Exploratory analyses included comparison of immunogenicity outcomes to adolescent (aged 12 to <18 years) participants from the 2019nCoV-301 phase 3 study. This comparator group had received two study doses of the prototype vaccine, NVX-CoV2373 (21 days apart), as a primary series and a 3^rd^ dose within 5 months of completion of the primary series. Other exploratory analyses were immunogenicity responses to the JN.1 strain. These assessments were conducted in a subset (90 baseline anti-nucleoprotein [NP] positive and ∼10 baseline anti-NP negative) of participants; the anti-NP baseline seropositive participants had received at least three prior vaccine doses.

### Ethics approval

The trial protocol was approved by the Independent Ethics Committee and the study was performed in accordance with the Declaration of Helsinki and the International Conference on Harmonization Good Clinical Practice guidelines. Clinical monitoring was conducted by Syneos Health (Morrisville, NC).

## Results

Of 433 participants screened for eligibility from September 7–26, 2023, 401 (93%) were randomised, and 32 (7%) were excluded; 24 (6%) did not meet inclusion/exclusion criteria, two (<1%) withdrew consent, and six (1%) were not randomised prior to enrolment closure (Figure 1). Of the 401 randomised participants, 190 (47%) participants were randomised to receive NVX-CoV2601, and 211 (53%) participants were randomised to receive the bivalent vaccine. The safety analysis sets included all participants randomised to the NVX-CoV2601 group and 210/211 (<100%) participants in the bivalent group (one vaccine was not administered). The day-28 per-protocol analysis sets (database lock 17 May 2024) included 178/190 (94%) and 194/211 (92%) participants in the NVX-CoV2601 and bivalent vaccine groups, respectively.

**Figure 1.**
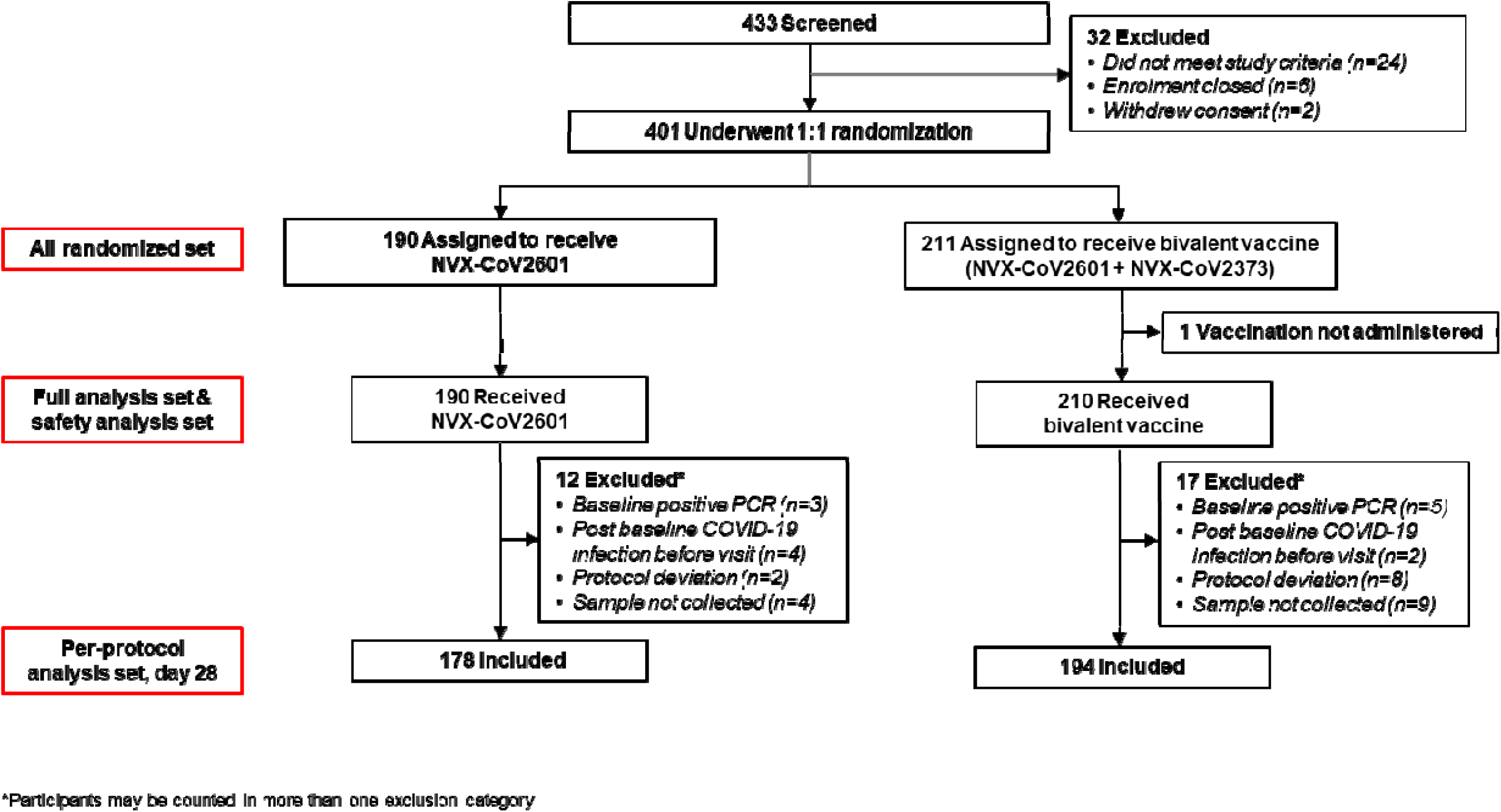
CONSORT diagram.

Participant demographics were balanced between the day-28 per-protocol analysis sets (Table 1). Overall, there were slightly more female (194/372 [52%]) than male participants, and most participants were White (269/372 [72%]). Median age (interquartile range [IQR]) in the NVX-CoV2373 and bivalent vaccine groups was 15.0 years (13.0–16.0) and 14.5 years (13.0– 16.0), respectively. Most participants had received two prior (NVX-CoV2601: 77/178 [43%]; bivalent: 89/194 [46%]) or three prior (67/178 [38%]; 75/194 [39%]) mRNA COVID-19 vaccines (versus four or five prior vaccines). Median days (IQR) since the most recent dose to study vaccination were 585.5 (340.0–658.0) and 593.5 (369.0–708.0) for NVX-CoV2601 and the bivalent vaccine, respectively. Demographics in the safety analysis sets were comparable between the groups and followed the same trends as in the per-protocol analysis sets (Table S1).

**Table 1.** Demographics and baseline clinical characteristics in the day-28 per-protocol analysis sets.

| Characteristic | NVX-CoV2601<br>(N=178) | Bivalent vaccine<br>(N=194) | NVX-CoV2373*<br>(N=114) |
| --- | --- | --- | --- |
| **Age, years** |  |  |  |
| Mean (SD) | 14.6 (1.73) | 14.5 (1.67) | 13.9 (1.47) |
| Median (IQR) | 15.0 (13–16) | 14.5 (13–16) | 14.0 (12–15) |
| **Sex** |  |  |  |
| Female | 95 (53.4) | 99 (51.0) | 47 (41.2) |
| Male | 83 (46.6) | 95 (49.0) | 67 (58.8) |
| **Race** |  |  |  |
| White | 128 (71.9) | 141 (72.7) | 103 (90.4) |
| Black or African American | 30 (16.9) | 37 (19.1) | 3 (2.6) |
| Multiple | 9 (5.1) | 10 (5.2) | 6 (5.3) |
| Asian | 8 (4.5) | 2 (1.0) | 2 (1.8) |
| Other | 2 (1.1) | 2 (1.0) | 0 |
| Native Hawaiian/<br>Other Pacific Islander | 1 (0.6) | 1 (0.5) | 0 |
| Not reported | 0 | 1 (0.5) | 0 |
| **Ethnicity** |  |  |  |
| Not Hispanic or Latino | 136 (76.4) | 151 (77.8) | 96 (84.2) |
| Hispanic or Latino | 42 (23.6) | 43 (22.2) | 18 (15.8) |
| **BMI (kg/m<sup>2</sup>)** |  |  |  |
| Underweight (<5th percentile) | 2 (1.1) | 4 (2.1) | 7 (6.1) |
| Normal (5<sup>th</sup>–<85<sup>th</sup> percentile) | 95 (53.4) | 108 (55.7) | 69 (60.5) |
| Overweight (85<sup>th</sup>–<95<sup>th</sup> percentile) | 37 (20.8) | 31 (16.0) | 10 (8.8) |
| Obese (≥95<sup>th</sup> percentile) | 44 (24.7) | 51 (26.3) | 28 (24.6) |
| **Prior mRNA-based COVID-19 vaccinations** |  |  |  |
| 2 doses | 77 (43.3) | 89 (45.9) | NA |
| 3 doses | 67 (37.6) | 75 (38.7) | NA |
| 4 doses | 33 (18.5) | 29 (14.9) | NA |
| 5 doses | 1 (0.6) | 1 (0.5) | NA |
| **Days since most recent prior COVID-19 vaccine** |  |  |  |
| Mean (SD) | 530.5 (191.21) | 550.5 (192.07) | – |
| Median (IQR) | 585.5 (340.0–658.0) | 593.5 (369.0–708.0) | – |
| **Baseline anti-N/PCR** |  |  |  |
| Positive | 167 (93.8) | 185 (95.4) | 0 |
| Negative | 11 (6.2) | 9 (4.6) | 114 (100) |

Characteristics are displayed as n (%), unless otherwise noted.
BMI=body mass index; IQR=interquartile range; NA=not applicable; PCR=polymerase chain reaction; SD=standard deviation.
\*The 2019nCoV-301 phase 3 study included a paediatric expansion group of adolescents aged 12 to <18 years who received two primary series doses of NVX-CoV2373 and a 3rd dose within 5 months of the primary series.

Any solicited TEAE occurred in 153/190 (81%) and 166/210 (79%) participants in the NVX-CoV2601 and bivalent safety analysis sets, respectively, the majority of which were grade 1/2 (grade 3: 3 [2%] and 5 [2%]; Table 2); there were no solicited events grade >3. Overall, solicited TEAEs (local or systemic) had a median duration of 2 days. Any solicited local event occurred in 136/190 (72%) participants who received NVX-CoV2601 and in 140/210 (67%) participants who received the bivalent vaccine (Figure 2A; Table 2). Grade 3 events occurred in ≤1% of participants in each group. Tenderness and pain were the most common solicited local reactions (>5% in either group), occurring in 112/190 (59%) and 99/190 (52%) participants in the NVX-CoV2601 group, respectively, and in 116/210 (55%) and 96/210 (46%) participants in the bivalent vaccine group, respectively. There were 4/190 (2%) participants in the NVX-CoV2601 group who collectively reported a total of six solicited local TEAEs that lasted >7 days post vaccination (tenderness=3; pain=2; swelling=1); 1/210 (<1%) participants in the bivalent group reported a TEAE (tenderness) lasting >7 days.

**Figure 2.**
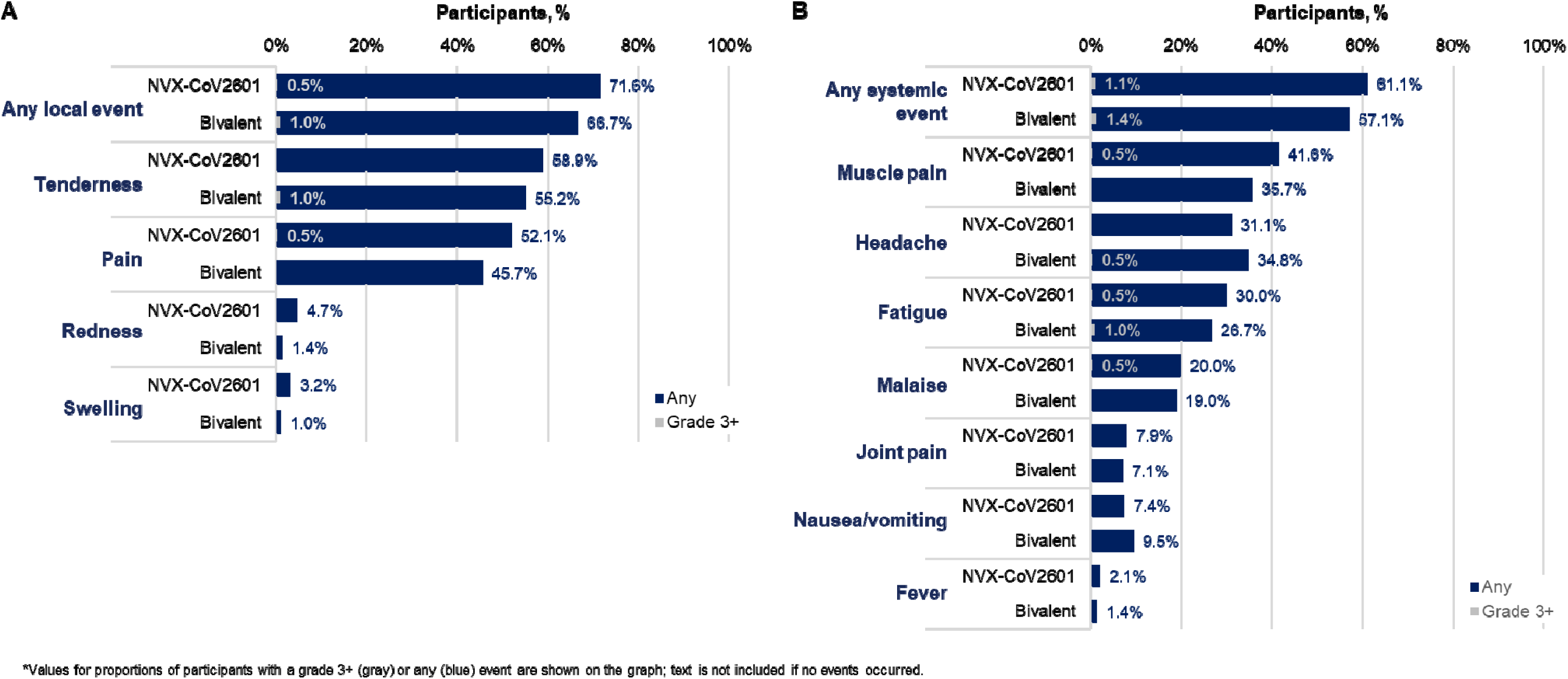
Proportion of participants with a solicited (A) local or (B) systemic reactogenicity event. The proportion of participants with a solicited treatment-emergent adverse event in the safety analysis sets for the NVX-CoV2601 and bivalent vaccine groups are shown for (A) local and (B) systemic reactogenicity. Percentages for any-grade events are shown in blue at the top of each bar and for grade ≥3 events in gray within the bars.

**Table 2.**
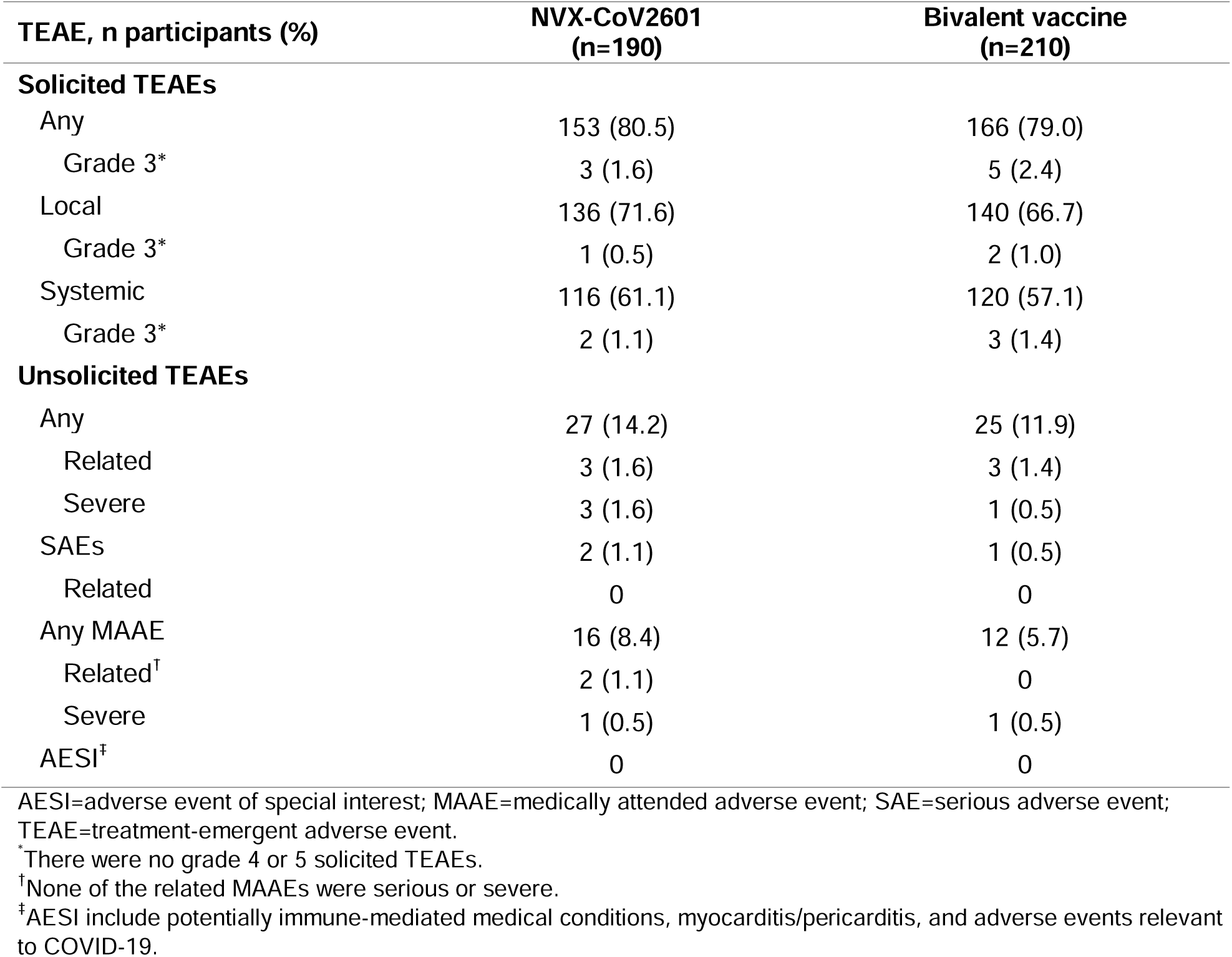
Safety summary of solicited and unsolicited TEAEs in the safety analysis sets.

Any solicited systemic event occurred in 116/190 (61%) and 120/210 (57%) participants in the NVX-CoV2601 and bivalent vaccine groups (grade 3: 2 [1%] and 3 [1%]), respectively (Figure 2B; Table 2). The most common (occurring in >20% of participants in either group) systemic TEAEs were muscle pain, headache, and fatigue. Of 190 participants in the NVX-CoV2601 group, four (2%) collectively reported a total of six solicited systemic TEAEs that lasted >7 days post vaccination (joint pain=2; fatigue=1; headache=1; malaise=1; nausea/vomiting=1). Solicited systemic reactions lasting >7 days were reported by 3/210 (1%) participants in the bivalent group (fatigue=3; headache=2; nausea/vomiting=1).

Throughout the study, unsolicited TEAEs occurred in 27/190 (14%) participants in the NVX-CoV2601 group and 25/210 (12%) participants in the bivalent group (Table 2). Almost all unsolicited events were considered to be unrelated to vaccination as well as mild-to-moderate in severity; related TEAEs occurred in 3/190 (2%) and 3/210 (1%) participants in the NVX-CoV2601 and bivalent vaccine groups, respectively. Severe events were experienced by three participants in the NVX-CoV2601 group (psychiatric disorder, n=2; infection/infestation, n=1) and one participant in the bivalent group (psychiatric disorder); none were related to the respective study vaccines. SAEs occurred in 4/190 (2%) and 1/210 (<1%) participants in the NVX-CoV2601 and bivalent groups, respectively; these were each psychiatric disorders and considered unrelated to study vaccination. MAAEs occurred in 16/190 (8%) participants in the NVX-CoV2601 group and in 12/210 (6%) participants in the bivalent group. Three MAAEs related to NVX-CoV2601 (injection site induration, pain in extremity, and urticaria) occurred among two participants; there were no vaccine-related MAAEs in the bivalent group. No TEAEs led to study discontinuation, and there were no AESIs (including no PIMMCs) or myocarditis/pericarditis in either group.

Neutralising antibody GMTs (ID_50_) to XBB.1.5 on day 0 were 208 in the NVX-CoV2601 group and 185 in the bivalent group (Figure 3A; Table S2). Respectively, titres were elevated 12.2-fold (95% CI 9.5–15.5) and 8.4-fold (95% CI 6.8–10.3) on day 28 post vaccination. Between-group day-28 GMTR (95% CI) indicates a lower response with the bivalent vaccine compared to NVX-CoV2601 (GMTR: 0.6 [95% CI 0.50–0.79]), and SRRs demonstrated a similar trend (Table S2). Responses against ancestral SARS-CoV-2 from day 0 to day 28 were elevated in both the NVX-CoV2601 (GMFR: 2.7 [95% CI 2.3–3.1]) and bivalent (GMFR: 2.3 [95% CI 2.0–2.7]) groups (Figure 3A; Table S2). The GMTR (0.8 [95% CI 0.7–1.0]) and SRR difference (−4.9% [95% CI −14.4 to 4.6]) against the ancestral virus indicated a more robust response in the NVX-CoV2601 group compared with the bivalent group (Table S2). Responses against XBB.1.5 were durable over time for both NVX-CoV2601 and the bivalent vaccine, with respective day-180 titres remaining 6.0-fold (95% CI 4.7–7.8) and 4.7-fold (95% CI 3.7–5.9) above baseline (GMT [95% CI]—NVX-CoV2601: 1370 [1128.3–1662.5]; bivalent: 866 [733.5–1022.7]) (Table S2; Figure S2).

**Figure 3.**
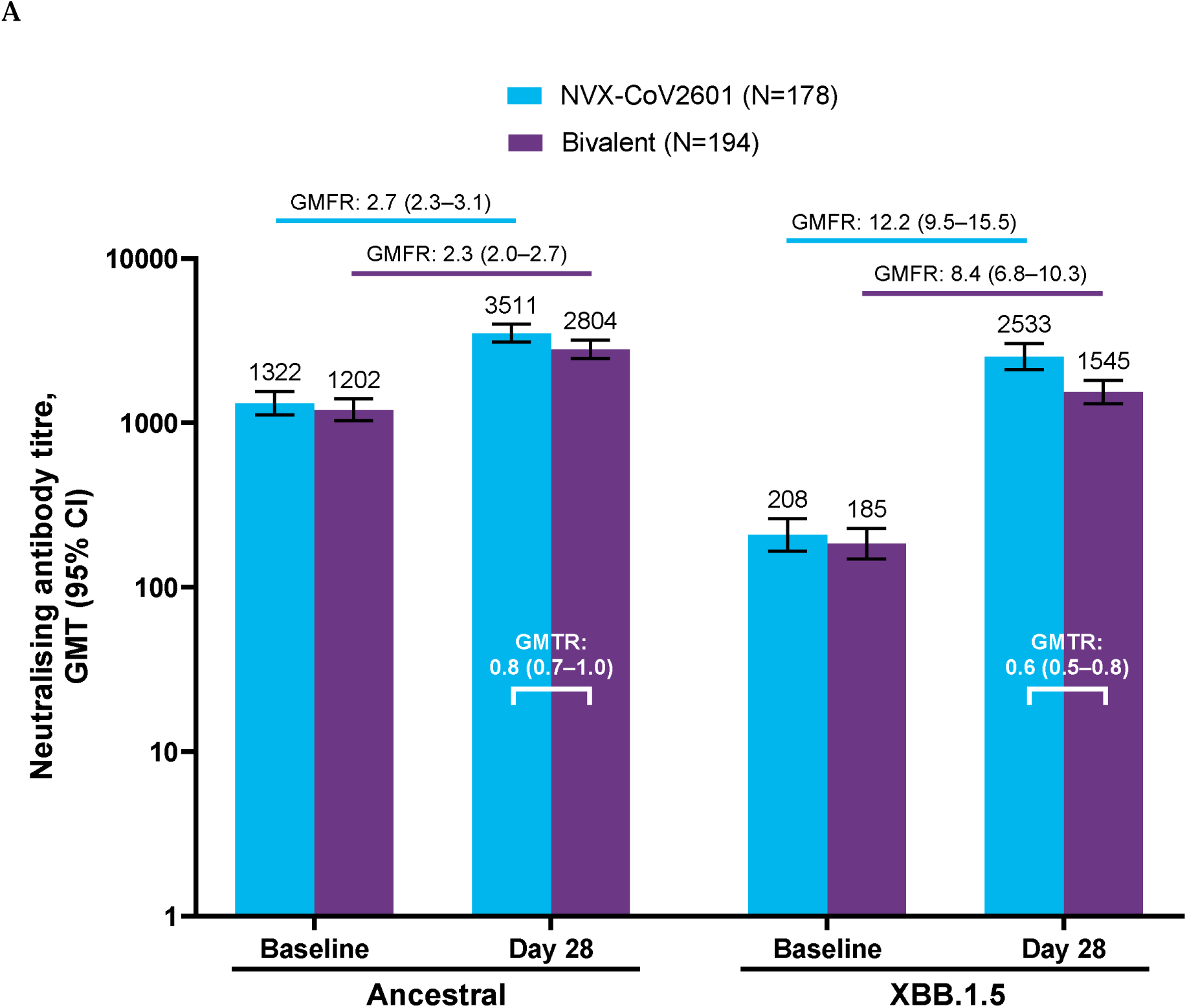
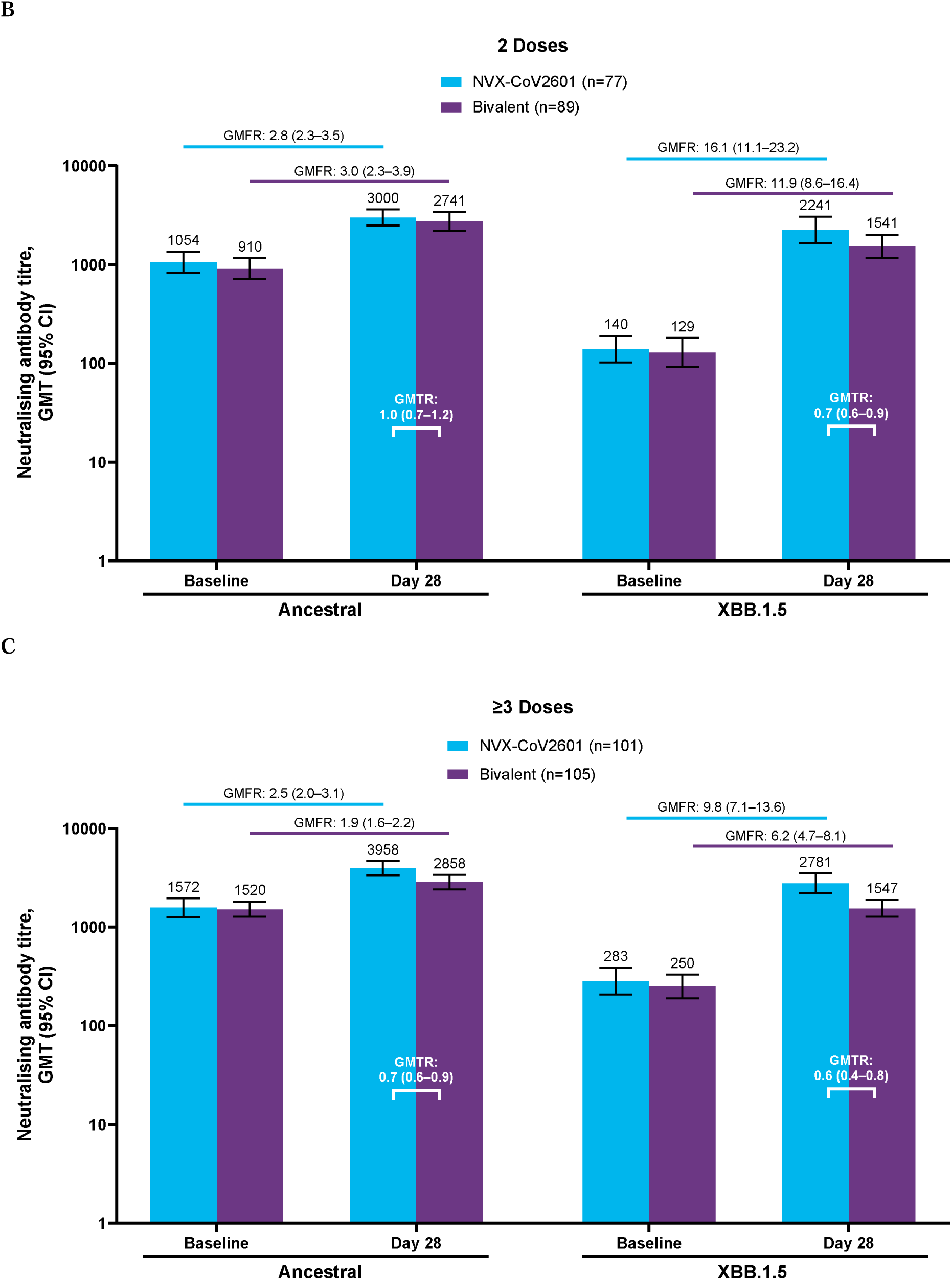
Pseudovirus neutralising antibody responses against ancestral and XBB.1.5 SARS-CoV-2 (A) overall and (B) by number of prior COVID-19 vaccinations. Participants were in subgroups of two or ≥3 prior doses of an mRNA-based vaccine (BNT162b2 and/or mRNA-1273) GMTs of neutralising antibody responses (ID_50_) to the ancestral virus or XBB.1.5 are shown on a log scale *y*-axis for (A) all participants and (B) whether two or ≥3 prior doses of an mRNA-based vaccine had been received. Corresponding GMFRs comparing baseline and day 28 GMTs are shown above each bar. GMFR=geometric mean fold rise; GMT=geometric mean titre; nAb=neutralising antibody.

For both strains, increases in neutralising antibodies were observed in a subgroup analysis of participants who received either two or ≥3 prior mRNA-based COVID-19 vaccines (Figure 3B; Table S3). Baseline and day-28 titres were comparable to the overall population; however, baseline GMTs were higher in those who received ≥3 versus two prior vaccinations. GMFRs from baseline indicated a more robust response in the 2-dose versus ≥3-dose groups for both the ancestral and XBB.1.5 strains.

In an exploratory immunogenicity analysis, pseudovirus neutralising antibody responses to the JN.1 strain were assessed in a subset of ∼100 participants in each of the two study vaccine groups. From comparable baseline titres, GMTs increased in both the NVX-CoV2601 and bivalent vaccine groups (Figure S3; Table S4). From day 0 to day 28, there was a more robust cross-reactive response (GMFR [95% CI]) to JN.1 by NVX-CoV2601 (11.1 [8.3–14.8]) than the bivalent vaccine (8.9 [7.0–11.2]); SRRs at day 28 were comparable.

Anti-rS IgG responses (GMEU) to XBB.1.5 rose from 38,994 at baseline to 150,233 at day 28 in the NVX-CoV2601 group and from 32,857 at baseline to 113,032 at day 28 in the bivalent group (Figure S4; Table S5). The two groups had comparable GMFRs (NVX-CoV2601: 3.9 [95% CI 3.3–4.4], bivalent: 3.4 [3.0–3.9]; Table S4). GMEUR (0.8 (95% CI 0.7–0.9]) and SRR differences (−8.5% ([95% CI −18.4 to 1.6]) suggest a stronger response with NVX-CoV2601 compared with the bivalent vaccine. Anti-rS IgG responses (GMEU) to ancestral SARS-CoV-2 were an exploratory objective. A baseline value of 71,535 increased to 181,737 at day 28 in the NVX-CoV2601 group; responses increased from 61,443 at baseline to 157,078 at day 28 in the bivalent group (Figure S4; Table S5). GMFRs and SRRs against ancestral SARS-CoV-2 were slightly lower than observed against XBB.1.5 for both vaccine groups but were comparable to each other. Responses against XBB.1.5 were durable for both NVX-CoV2601 and the bivalent vaccine, with day-180 titres for both vaccines at 1.9-fold (95% CI 1.7–2.2) above baseline (day-28 GMEUs [95% CI]—NVX-CoV2601: 77,150 [68,110.5–87,388.8]; bivalent: 61,648 [55,459.2–68,526.7]; Table S5). In a subgroup analysis of participants by number of prior COVID-19 vaccinations, the highest titres were observed on day 28 in participants with ≥3 pre-study vaccinations (Figure S4; Table S6). Within this subset, the largest increase from baseline titre was observed against XBB.1.5 in participants with two prior vaccinations.

As part of the exploratory objective to describe neutralising antibody responses, GMTs (ID_50_) were assessed for the NVX-CoV2601 group compared with a representative adolescent population (aged 12 to <18 years) from the phase 3 2019nCoV-301 study (Table 1) who received a primary series and additional dose of the prototype vaccine, NVX-CoV2373. As stated earlier, the NVX-CoV2601 group had neutralising antibody titres increase from 208 to 2533 (baseline to day 28) against XBB.1.5 (Figure 3A; Table S2). Compared with the NVX-CoV2601 group, the NVX-CoV2373 comparator group had less of an increase from baseline (GMT: 20) to day 28 (GMT: 114; GMFR: 5.6 [95% CI 4.6–6.7]; Figure S5, Table S7). The GMTR (12.2 [95% CI 8.5–17.4]) and SRR difference (32.2% [95% CI 20.7–42.9]) between NVX-CoV2601 and the NVX-CoV2373 comparator group also reflects a more robust response to XBB.1.5 for the matched vaccine. Baseline titres against the ancestral strain were higher for NVX-CoV2601 compared with the NVX-CoV2373 comparator (1322 vs 202); however, both groups produced comparable day-28 titres (3511 vs 4200; Figure S5, Table S7). Both NVX-CoV2601 and NVX-CoV2373 responded to ancestral SARS-CoV-2; however, the NVX-CoV2373 comparator group had a more robust response to its matched, ancestral SARS-CoV-2 strain (GMTR: 0.6 [0.44–0.71]) than that observed for NVX-CoV2601.

## Discussion

A tolerable safety profile and durable immunogenicity were demonstrated for NVX-CoV2601 (an XBB.1.5-directed protein vaccine) in adolescents when given as a heterologous dose following prior vaccination with mRNA-based (mRNA-1273 and/or BNT162b2) COVID-19 vaccines. Robust antibody responses were observed in both vaccine groups; however, the monovalent formulation generated higher day-28 neutralising antibody titres to the XBB.1.5 pseudovirus compared with a bivalent version of the vaccine containing the rS for XBB.1.5 and ancestral SARS-CoV-2. The safety profiles of both the monovalent and bivalent variant-specific vaccines were consistent with the established safety profile of the prototype vaccine in adults and adolescents^2,25^ and of NVX-CoV2601 in adults.^20^

Notably, NVX-CoV2601 was able to increase antibody titres to ancestral SARS-CoV-2 (Wuhan strain) to a greater degree than the bivalent vaccine (GMFR: 2.7 vs 2.3, respectively), even though it did not contain the rS to the ancestral antigen. Neutralising antibody and anti-rS IgG responses against XBB.1.5 by NVX-CoV2601 were also more pronounced than those with the bivalent vaccine when analysed in participant subsets based on the number of prior mRNA-based COVID-19 vaccines (2 prior doses: GMFR 16.1 vs 11.9, respectively; ≥3 prior doses: GMFR 9.8 vs 6.2, respectively). A more pronounced response with NVX-CoV2601 versus the bivalent vaccine may reflect the effects of prior immune imprinting to ancestral SARS-CoV-2 combined with the fact that monovalent NVX-CoV2601 contains 5 µg of a single type of rS whereas the bivalent vaccine contains a half dose (2.5 µg) of two different rS proteins, indicating that a full dose of rS may be preferred compared with a half dose. A similar pattern was seen with mRNA vaccines, in which the Omicron BA.1 monovalent vaccines elicited more robust immune responses compared with the bivalent vaccine (prototype + Omicron BA.1).^26,27^ This is further supported by the observed responses of a full dose of rS against a heterologous virus (eg, NVX-CoV2601 against the ancestral virus and NVX-CoV2373 against XBB.1.5) in the 2019nCoV-301 study comparator analysis. When assessed against a comparator group of adolescents who received prototype NVX-CoV2373 during the 2019nCoV-301 study, both groups had an increase in neutralising antibodies from baseline; however, as anticipated, NVX-CoV2601 demonstrated more robust neutralising responses against the XBB.1.5 pseudovirus. NVX-CoV2601 produced immune responses against the ancestral strain as well, although these were lower than those it produced against XBB.1.5 and lower compared with responses of NVX-CoV2373 against the ancestral strain. These results indicate some degree of cross-neutralising activity and provide supportive evidence for the adaptability of this vaccine platform to address COVID-19 vaccine formulation updates that are recommended for alignment with changes in predominantly circulating variants. Although the bivalent formulation used in this study has not been authorised or approved for use, NVX-CoV2601 was authorised in the US, the European Union, and the United Kingdom for the 2023–2024 season.

A limitation of the present study is that it was solely conducted in a US population, and the majority (∼70%) of participants were White; however, there are no indications that immunogenicity to COVID-19 vaccines varies based on race^28^. Additionally, this study was not designed to assess vaccine efficacy, nor was it powered to make formal statistical statements on immunogenicity or to detect rare adverse events such as myocarditis. It is important to note that the dynamics of the pandemic were very different when participants were enroled into the pivotal 2019nCoV-301 study (December 2020 to February 2021)^2^, during which natural exposure was relatively low, compared with enrolment for the current study (September 2023), when natural infection or prior vaccination is much more prevalent. As such, the results should be cautiously interpreted with those caveats in mind since prior exposure and/or vaccination could prime the immune system (as indicated in this study by the baseline titres), making it possible to mount a more robust response to subsequent virus exposures. Furthermore, blood samples for immunogenicity assessments were largely collected when XBB.1.5 and XBB.1.16 variants were predominant. Finally, this study did not include a placebo control group; however, this study design is typical of clinical trials investigating updated vaccines,^26,27,29,30^ and it is difficult to include unvaccinated controls since SARS-CoV-2 vaccines are recommended by regulatory authorities in this age group^31^.

In July 2024, the JN.1-lineage descendants KP.2 and KP.3 emerged as the most prevalent SARS-CoV-2 strains in the US, each having developed three additional spike gene mutations from the JN.1 sequence, which could potentially further increase their immune evasion capabilities.^27,32^ The exploratory analyses described here demonstrated a cross-reactive response to the JN.1 strain by both the XBB.1.5 and the bivalent vaccines. A similar cross-reactive response was seen in adults for JN.1 and the KP.2 subvariant.^20^ While data have yet to be gathered regarding the efficacy of updated JN.1-targeted vaccines against substrains like KP.2 and KP.3, preliminary preclinical data suggest strong cross-reactive immune responses. For example, sera from individuals with COVID-19 when JN.1 was predominant (n=7; November 2023 to February 2024) effectively neutralised KP.2 and other JN.1 subvariants.^33^ While neutralisation of KP.3 was not investigated, preclinical data suggest no significant difference in pseudovirus neutralisation resistance between KP.2 and KP.3.^34^ Finally, pseudovirus neutralisation of KP.2 (and other variants) was observed in mice and nonhuman primates after boosting with a formulation of the nanoparticle protein vaccine with Matrix-M™ adjuvant that contains rS for JN.1.^15^

These results provide clinical data to support that the updated vaccine, NVX-CoV2601, containing XBB.1.5 rS is safe in adolescents aged 12 to <18 years and provides durable immunogenicity against more than one SARS-CoV-2 strain. These findings reinforce recommendations to receive updated variant-based vaccines for the 2024-2025 season.

## Author contributions

CB and RMM were involved in the study design. EC, LC, JiA, ABH, and JMA were involved in data collection. CB and GC accessed and verified the data. GC performed the statistical analyses. CB, GC, EC, LC, JiA, ABH, JSP, MZ, SCC, ZC, RK, KH, KS, SN, FN, RMM, and JMA were involved in data interpretation and reviewed, commented on, and approved this manuscript prior to submission for publication. The authors were not precluded from accessing data in the study. The authors accept accountabilities for all aspects of the work, ensuring questions related to accuracy/integrity are investigated and resolved, and accept responsibility to submit the manuscript for publication.

## Acknowledgments

We thank Jibran Atwi, MD, of Velocity Clinical Research (Louisiana, USA), Matthew Braddock, DO, of Westside Center for Clinical Research (Florida, USA), Kenneth Etokahana, MD, of Tekton Research (Texas, USA), Douglas Logan, MD, of Velocity Clinical Research (Ohio, USA), and Divina Roman, MD, of Tekton Research (Oklahoma, USA), who are investigators in the 2019nCoV-314 Study and gave feedback on an earlier draft of the manuscript. We also thank all the study participants who volunteered for this study. This study was funded by Novavax, Inc. Medical writing and editorial support were provided by Kelly M. Fahrbach, PhD, CMPP, and Ebenezer M. Awuah-Yeboah, BS, of Ashfield MedComms (New York, USA), an Inizio company, supported by Novavax, Inc.

## Appendix A. Supplementary material

Supplementary tables and figures.

## Data availability

The study information is available at https://clinicaltrials.gov/study/NCT05973006. Requests submitted to the corresponding author will be considered upon publication of this article, and deidentified participant data may be provided. The study protocol is available in online supplemental material.

