## Supplementary material for "Safety and Immunogenicity of Omicron Protein Vaccines in mRNA-Vaccinated Adolescents: A Phase 3, Randomised Trial": N/A

**Table of contents**

**Figure S1.** Study visits and sample collection	3

**Figure S2. N**eutralising antibody titres to ancestral SARS-CoV-2 or XBB.1.5 over time	4

**Figure S3.** Pseudovirus neutralising antibody responses against JN.1 SARS-CoV-2	5

**Figure S4.** Anti-rS IgG responses against ancestral and XBB.1.5 SARS-CoV-2 (A) overall and (B) by number of prior COVID-19 vaccinations	6

**Figure S5.** Neutralising antibody responses to ancestral and XBB.1.5 SARS-CoV-2 for NVX-CoV2601 and an NVX-CoV2373 comparator group	8

**Table S1.** Demographics and baseline clinical characteristics in the full and safety analysis sets	9

**Table S2.** Pseudovirus neutralising antibody responses against ancestral and XBB.1.5 SARS-CoV-2 (per-protocol analysis sets)	10

**Table S3.** Pseudovirus neutralising antibody responses against ancestral and XBB.1.5 SARS-CoV-2 by number of prior COVID-19 vaccines (per-protocol analysis sets)	11

**Table S4.** Pseudovirus neutralising antibody responses against JN.1 SARS-CoV-2 (per-protocol analysis subset).12

**Table S5.** Anti-spike IgG responses against ancestral and XBB.1.5 SARS-CoV-2 strains	13

**Table S6.** Anti-rS IgG responses against ancestral and XBB.1.5 SARS-CoV-2 strains by number of prior COVID-19 vaccinations (per-protocol analysis sets)	14

**Table S7.** Pseudovirus neutralising antibody response comparison of NVX-CoV2601 and NVX-CoV2373 (per-protocol analysis sets)	15

**2019nCoV-314 Study Investigators**	16

**Figure S1.** Study visits and sample collection

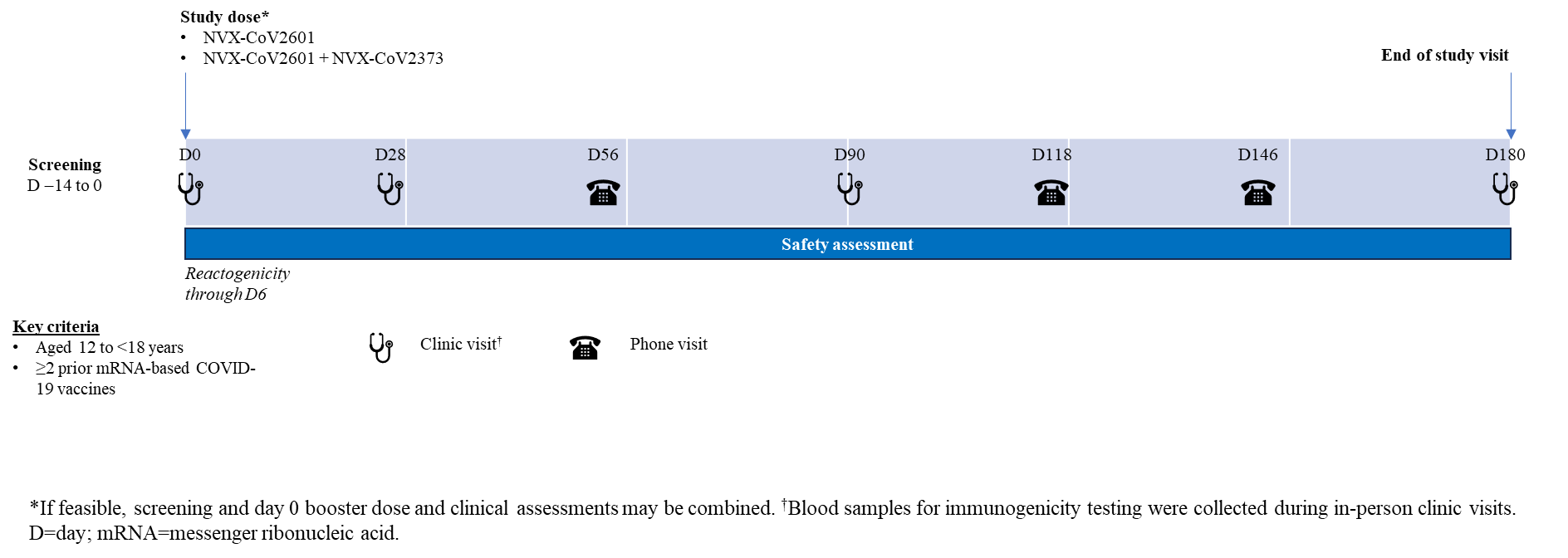

**Figure S2. Neutralising antibody titres to ancestral SARS-CoV-2 or XBB.1.5 over time**

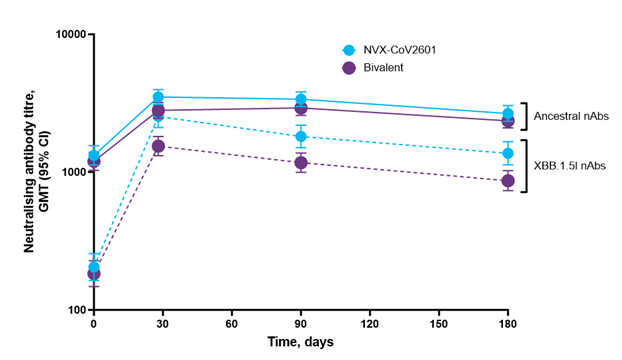

**Figure S3.** Pseudovirus neutralising antibody responses against JN.1 SARS-CoV-2

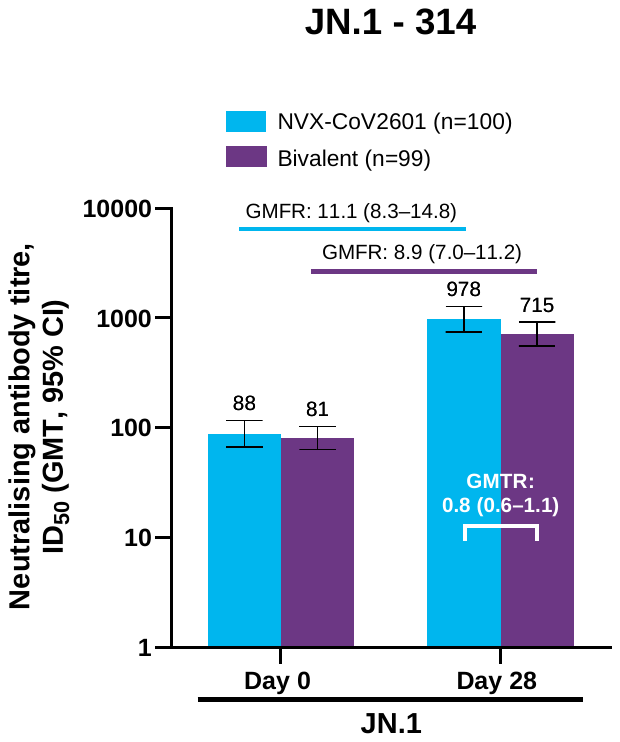

**Figure S4.** Anti-rS IgG responses against ancestral and XBB.1.5 SARS-CoV-2 (A) overall and (B) by number of prior COVID-19 vaccinations

**A**

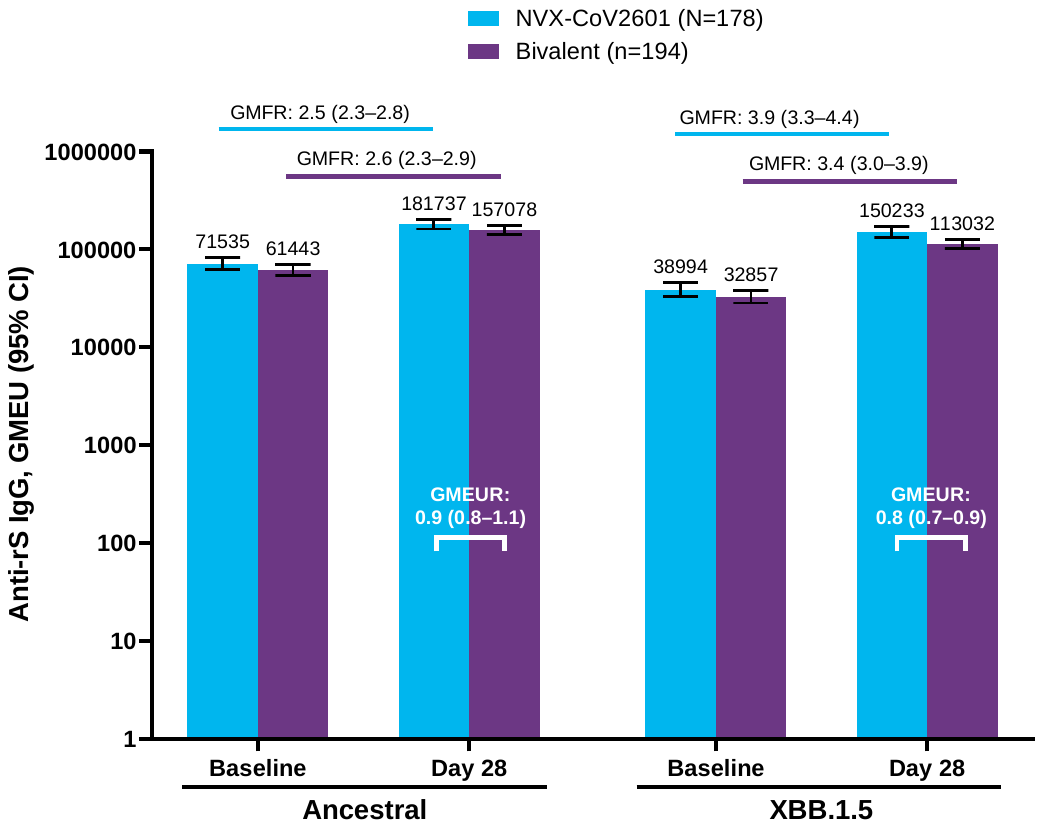

**B**

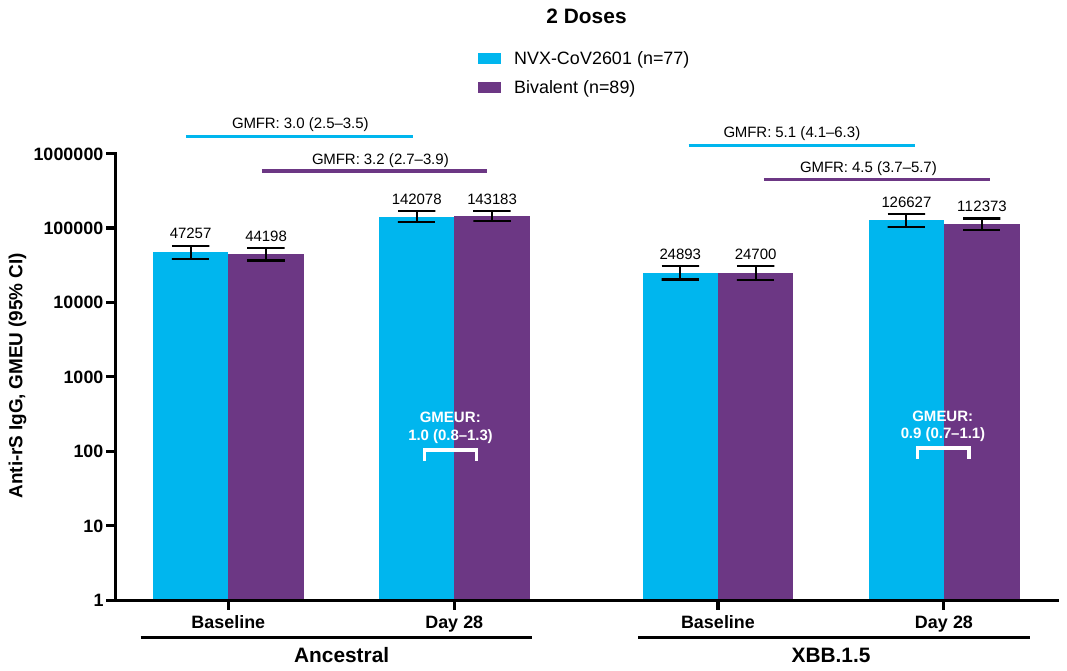

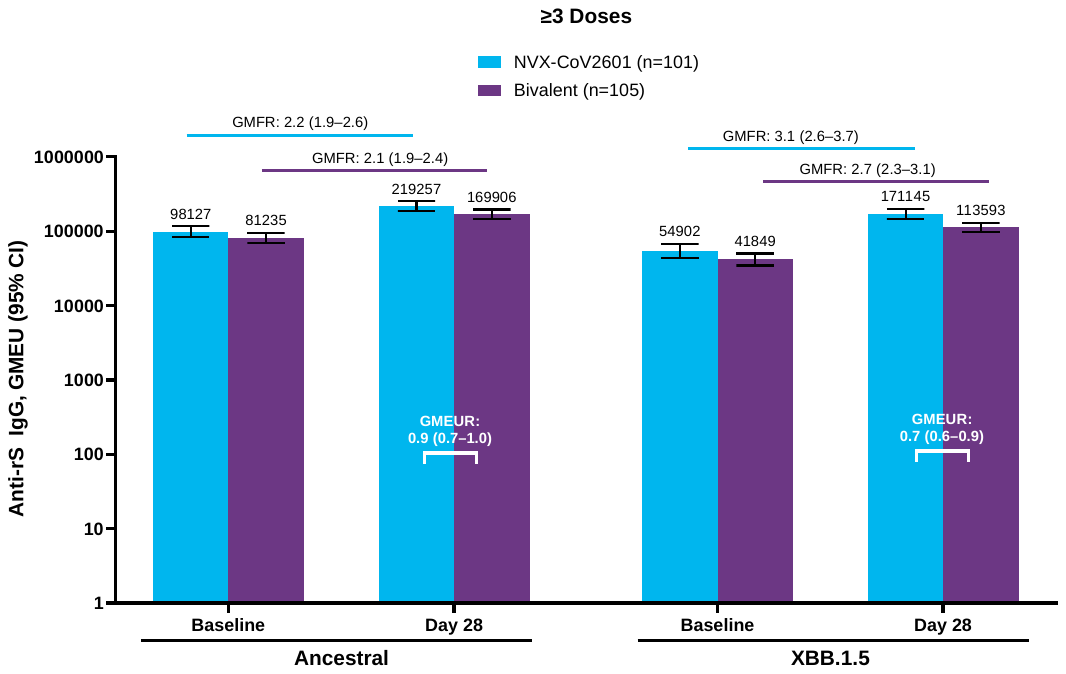

**Figure S5.** Neutralising antibody responses to ancestral and XBB.1.5 SARS-CoV-2 for NVX-CoV2601 and an NVX-CoV2373 comparator group

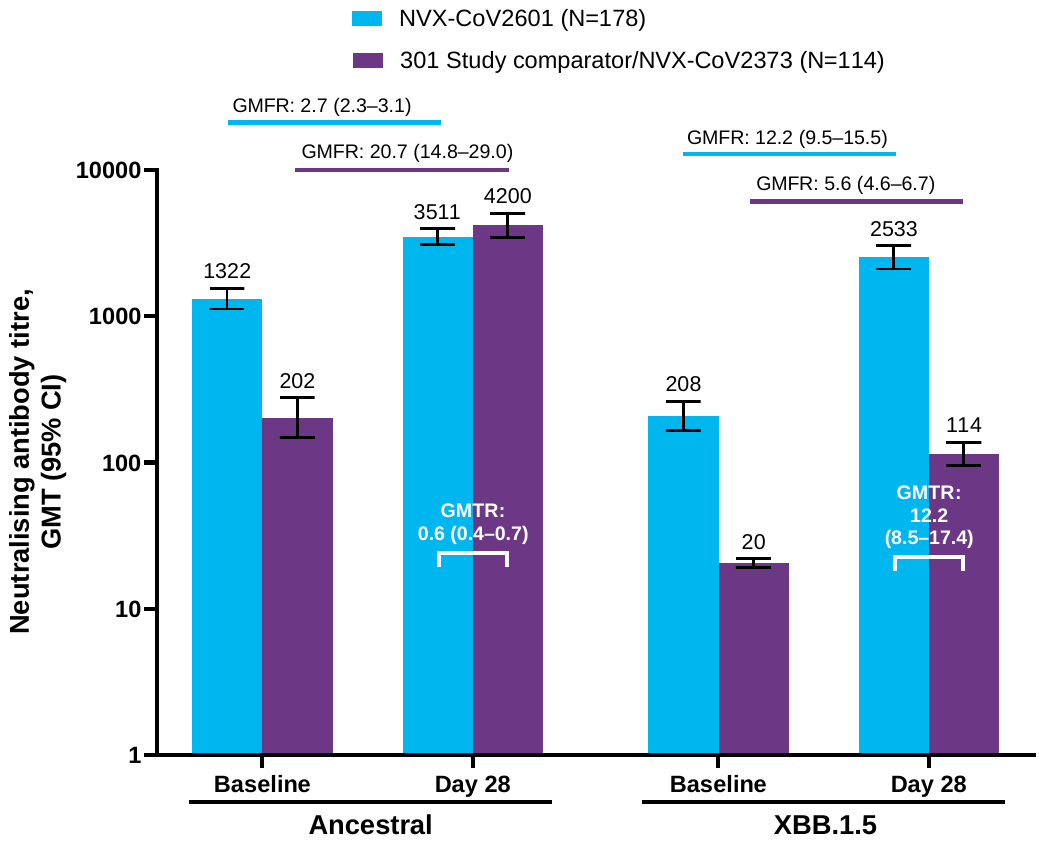

**Table S1.** Demographics and baseline clinical characteristics in the full and safety analysis sets

| Characteristic | NVX-CoV2601  (N=190) | Bivalent vaccine  (N=210) |
| --- | --- | --- |
| Age, years |  |  |
| Mean (SD) | 14.5 (1.77) | 14.5 (1.65) |
| Median (IQR) | 15.0 (13.0–16.0) | 14.0 (13.0–16.0) |
| Sex |  |  |
| Female | 99 (52.1) | 107 (51.0) |
| Male | 91 (47.9) | 103 (49.0) |
| Race |  |  |
| White | 137 (72.1) | 149 (71.0) |
| Black or African American | 33 (17.4) | 44 (21.0) |
| Multiple | 9 (4.7) | 10 (4.8) |
| Asian | 8 (4.2) | 2 (1.0) |
| Other | 2 (1.1) | 2 (1.0) |
| Native Hawaiian/ Other Pacific Islander | 1 (0.5) | 1 (0.5) |
| Unknown/not reported | 0 | 2 (1.0) |
| Ethnicity |  |  |
| Not Hispanic or Latino | 147 (77.4) | 163 (77.6) |
| Hispanic or Latino | 43 (22.6) | 47 (22.4) |
| BMI (kg/m^2^) |  |  |
| Underweight (<5th percentile) | 2 (1.1) | 4 (1.9) |
| Normal (5^th^−<85^th^ percentile) | 105 (55.3) | 120 (57.1) |
| Overweight (85^th^−<95^th^ percentile) | 38 (20.0) | 32 (15.2) |
| Obese (≥95^th^ percentile) | 45 (23.7) | 54 (25.7) |
| Prior COVID-19 vaccinations |  |  |
| 2 doses | 80 (42.1) | 96 (45.7) |
| 3 doses | 70 (36.8) | 82 (39.0) |
| 4 doses | 39 (20.5) | 31 (14.8) |
| 5 doses | 1 (0.5) | 1 (0.5) |
| Days since most recent prior COVID-19 vaccine |  |  |
| Mean (SD) | 522.5 (190.57) | 550.1 (191.08) |
| Median (IQR) | 574.0 (335.0–653.0) | 592.0 (364.0–708.0) |
| Baseline anti-N/PCR |  |  |
| Positive | 175 (92.1) | 200 (95.2) |
| Negative | 15 (7.9) | 10 (4.8) |

Characteristics are displayed as n (%), unless otherwise noted.

BMI=body mass index; IQR=interquartile range; NA=not applicable; SD=standard deviation.

**Table S2.** Pseudovirus neutralising antibody responses against ancestral and XBB.1.5 SARS-CoV-2 (per-protocol analysis sets)

| Parameter |  |  |
| --- | --- | --- |
| Day 28 per-protocol set | **NVX-CoV2601**  **(n=178)** | **Bivalent**  **(n=194)** |
| Ancestral |  |  |
| Day 0 GMT (95% CI) | 1322.20 (1122.43–1557.54) | 1201.54 (1033.73–1396.60) |
| Day 28 GMT (95% CI) | 3510.84 (3099.99–3976.14) | 2803.80 (2453.96–3203.51) |
| Day 28 adjusted GMT* (95% CI) | 3447.27 (3062.30–3880.62) | 2851.21 (2545.48–3193.65) |
| GMTR* between groups (95% CI) |  | 0.8 (0.70–0.97) |
| GMFR from baseline to day 28 (95% CI) | 2.7 (2.3–3.1) | 2.3 (2.0–2.7) |
| SRR^†^ from baseline to day 28 (95% CI) | 34.8% (27.9–42.3) | 29.9% (23.5–36.9) |
| SRR difference between groups (95% CI) |  | −4.9% (−14.4 to 4.6) |
| XBB.1.5 |  |  |
| Day 0 GMT (95% CI) | 208.40 (166.20–261.32) | 184.81 (148.86–229.45) |
| Day 28 GMT (95% CI) | 2533.08 (2107.17–3045.07) | 1544.61 (1314.33–1815.25) |
| Day 28 adjusted GMT* (95% CI) | 2489.35 (2112.17–2933.89) | 1569.49 (1340.94–1837.00) |
| GMTR* between groups (95% CI) |  | 0.6 (0.50–0.79) |
| GMFR from baseline to day 28 (95% CI) | 12.2 (9.5–15.5) | 8.4 (6.8–10.3) |
| SRR^†^ from baseline to day 28 (95% CI) | 70.8% (63.5–77.3) | 62.4% (55.1–69.2) |
| SRR difference between groups (95% CI) |  | −8.4% (−17.9 to 1.2) |
| Day 180 per-protocol set | **NVX-CoV2601 (n=180)** | **Bivalent (n=196)** |
| Ancestral |  |  |
| Day 0 GMT (95% CI) | 1321.00 (1123.04–1553.87) | 1197.59 (1029.90–1392.58) |
| Day 180 GMT (95% CI) | 2663.87 (2334.49–3039.72) | 2357.51 (2095.11–2652.77) |
| GMFR from baseline to day 180 (95% CI) | 1.9 (1.6–2.2) | 2.0 (1.7–2.3) |
| SRR from baseline to day 180 (95% CI) | 22.0 (16.0–29.1) | 26.6 (20.2–33.7) |
| XBB.1.5 |  |  |
| Day 0 GMT (95% CI) | 204.98 (163.68–256.69) | 183.37 (147.82–227.47) |
| Day 180 GMT (95% CI) | 1369.61 (1128.29–1662.53) | 866.10 (733.49–1022.67) |
| GMFR from baseline to day 180 (95% CI) | 6.0 (4.7–7.8) | 4.7 (3.7–5.9) |
| SRR from baseline to day 180 (95% CI) | 59.5% (51.7–67.0) | 49.7 (42.1–57.3) |

GMFR=geometric mean fold rise; GMT=geometric mean titre; GMTR=geometric mean titre ratio; LLOQ=lower limit of quantification; SRR=seroresponse rate.

*An analysis of covariance with vaccine group as fixed effect and baseline value as covariate is performed to estimate the adjusted GMT and GMTR. The mean difference between vaccine groups and the corresponding CI limits is then exponentiated to obtain the ratio of GMTs and the corresponding 95% CIs. Values <LLOQ are replaced by 0·5 $\times$ LLOQ for calculation of geometric means, GMFR, and GMTR.

^†^Seroresponse is defined as a ≥4-fold increase from baseline values ≥LLOQ or ≥4-fold the LLOQ if the baseline value is <LLOQ.

**Table S3.** Pseudovirus neutralising antibody responses against ancestral and XBB.1.5 SARS-CoV-2 by number of prior COVID-19 vaccines (per-protocol analysis sets)

| Parameter | NVX-CoV2601 | | | | Bivalent | | |
| --- | --- | --- | --- | --- | --- | --- | --- |
| Prior COVID-19 vaccines | **2 Doses**  **(n=77)** | **≥3 Doses**  **(n=101)** | | **2 Doses**  **(n=89)** | | | **≥3 Doses**  **(n=105)** |
| Ancestral |  | | |  | | | |
| Day 0 GMT (95% CI) | 1053.67 (823.74–1347.79) | 1572.04 (1265.56–1952.73) | | 910.28 (713.11–1161.98) | | | 1520.30 (1273.73–1814.60) |
| Day 28 GMT (95% CI) | 2999.79 (2490.49–3613.25) | 3958.19 (3352.12–4673.84) | | 2740.75 (2209.56–3399.63) | | | 2858.38 (2415.19–3382.89) |
| Day 28 adjusted GMT* (95% CI) | 2916.24 (2411.13–3527.17) | 3932.71 (3382.14–4572.91) | | 2808.56 (2353.22–3352.00) | | | 2876.19 (2480.71–3334.71) |
| GMTR* between groups (95% CI) | – | – | | 1.0 (0.74–1.25) | | | 0.7 (0.59–0.90) |
| GMFR from baseline to day 28 (95% CI) | 2.8 (2.3–3.5) | 2.5 (2.0–3.1) | | 3.0 (2.3–3.9) | | | 1.9 (1.6–2.2) |
| SRR^†^ from baseline to day 28 (95% CI) | 32.5 (22.2–44.1) | 36.6 (27.3–46.8) | | 41.6 (31.2–52.5) | | | 20.0 (12.8–28.9) |
| SRR difference between groups (95% CI) | – | – | | 9.1 (−5.7 to 23.4) | | | −16.6 (−28.6 to −4.4) |
| XBB.1.5 |  | |  | | |  | |
| Day 0 GMT (95% CI) | 139.50 (102.38–190.08) | 283.02 (206.98–386.98) | | 129.34 (92.66–180.55) | | | 250.10 (190.10–329.03) |
| Day 28 GMT (95% CI) | 2241.24 (1650.87–3042.72) | 2780.85 (2212.31–3495.50) | | 1541.31 (1180.82–2011.85) | | | 1547.42 (1268.02–1888.39) |
| Day 28 adjusted GMT* (95% CI) | 2211.58 (1683.46–2905.37) | 2737.52 (2236.01–3351.50) | | 1559.17 (1209.69–2009.62) | | | 1570.98 (1288.19–1915.85) |
| GMTR* between groups (95% CI) | – | – | | 0.7 (0.49–1.02) | | | 0.6 (0.43–0.76) |
| GMFR from baseline to day 28 (95% CI) | 16.1 (11.1–23.2) | 9.8 (7.1–13.6) | | 11.9 (8.6–16.4) | | | 6.2 (4.7–8.1) |
| SRR^†^ from baseline to day 28 (95% CI) | 76.6 (65.6–85.5) | 66.3 (56.2–75.4) | | 68.5 (57.8–78.0) | | | 57.1 (47.1–66.8) |
| SRR difference between groups (95% CI) | – | – | | −8.1 (−21.4 to 5.7) | | | −9.2 (−22.2 to 4.1) |

GMFR=geometric mean fold rise; GMT=geometric mean titre; GMTR=geometric mean titre ratio; LLOQ=lower limit of quantification; SRR=seroresponse rate.

*An analysis of covariance with vaccine group as fixed effect and baseline value as covariate is performed to estimate the adjusted GMT and GMTR. The mean difference between vaccine groups and the corresponding CI limits is then exponentiated to obtain the ratio of GMTs and the corresponding 95% CIs. Values <LLOQ are replaced by 0·5 $\times$ LLOQ for calculation of geometric means, GMFR and GMTR. ^†^Seroresponse is defined as a ≥4-fold increase from baseline values ≥LLOQ or ≥4-fold the LLOQ if the baseline value is <LLOQ.

**Table S4.** Pseudovirus neutralising antibody responses against JN.1 SARS-CoV-2 (per-protocol analysis subset*)

| Parameter | NVX-CoV2601  (n=100) | Bivalent  (n=99) |
| --- | --- | --- |
| JN.1 |  |  |
| Day 0 GMT (95% CI) | 88.1 (66.6–116.7) | 80.8 (63.4–103.0) |
| Day 28 GMT (95% CI) | 977.9 (749.7–1275.6) | 715.2 (555.4–921.0) |
| Day 28 adjusted GMT* (95% CI) | 957.3 (765.7–1196.9) | 730.8 (583.8–914.7) |
| GMTR* between groups (95% CI) |  | 0.8 (0.6–1.1) |
| GMFR from baseline to day 28 (95% CI) | 11.1 (8.3–14.8) | 8.9 (7.0–11.2) |
| SRR^†^ from baseline to day 28 (95% CI) | 75.0 (65.3–83.1) | 73.7 (63.9–82.1) |
| SRR difference between groups (95% CI) |  | −1.3 (−13.4 to 0.9) |

GMFR=geometric mean fold rise; GMT=geometric mean titre; GMTR=geometric mean titre ratio; LLOQ=lower limit of quantification; SRR=seroresponse rate.

*The subset for NVX-CoV2601 contained 90 baseline anti-NP positive and 10 baseline anti-NP negative participants; the subset for the bivalent vaccine group contained 90 and 9 participants, respectively. The anti-NP baseline seropositive participants had received at least three prior vaccine doses.

^†^An analysis of covariance with vaccine group as fixed effect and baseline value as covariate is performed to estimate the adjusted GMT and GMTR. The mean difference between vaccine groups and the corresponding CI limits is then exponentiated to obtain the ratio of GMTs and the corresponding 95% CIs. Values <LLOQ are replaced by 0·5 $\times$ LLOQ for calculation of geometric means, GMFR, and GMTR.

^†^Seroresponse is defined as a ≥4-fold increase from baseline values ≥LLOQ or ≥4-fold the LLOQ if the baseline value is <LLOQ.

**Table S5.** Anti-spike IgG responses against ancestral and XBB.1.5 SARS-CoV-2 strains

| Parameter |  |  |
| --- | --- | --- |
| Day 28 per-protocol set | **NVX-CoV2601**  **(n=178)** | **Bivalent**  **(n=194)** |
| Ancestral |  |  |
| Day 0 GMEU (95% CI) | 71,534.7 (62295.5–82144.1) | 61,442.8 (54028.4–69874.8) |
| Day 28 GMEU (95% CI) | 181,736.6 (162092.7–203761.1) | 157,077.8 (141109.2–174853.4) |
| Day 28 adjusted GMEU* (95% CI) | 174,493.5 (159641.9–190726.8) | 163,050.1 (149734.3–177549.9) |
| GMEUR* between groups (95% CI) |  | 0.9 (0.83–1.06) |
| GMFR from baseline to day 28 (95% CI) | 2.5 (2.3–2.8) | 2.6 (2.3–2.9) |
| SRR^†^ from baseline to day 28 (95% CI) | 27.5% (21.1–34.7) | 23.2% (17.5–29.8) |
| SRR difference between groups (95% CI) |  | −4.3% (−13.2 to 4.5) |
| XBB.1.5 |  |  |
| Day 0 GMEU (95% CI) | 38,993.6 (33248.9–45730.8) | 32,857.2 (28374.3–38048.3) |
| Day 28 GMEU (95% CI) | 150,232.9 (132853.6–169885.6) | 113,031.9 (101227.0–126213.4) |
| Day 28 adjusted GMEU* (95% CI) | 145,054.9 (131004.0–160612.9) | 116,728.6 (105876.4–128693.1) |
| GMEUR* between groups (95% CI) |  | 0.8 (0.70–0.93) |
| GMFR from baseline to day 28 (95% CI) | 3.9 (3.3–4.4) | 3.4 (3.0–3.9) |
| SRR^†^ from baseline to day 28 (95% CI) | 46.6% (39.1–54.2) | 38.1% (31.3–45.4) |
| SRR difference between groups (95% CI) |  | −8.5% (−18.4 to 1.6) |
| Day 180 per-protocol set | **NVX-CoV2601 (n=180)** | **Bivalent (n=196)** |
| Ancestral |  |  |
| Day 0 GMEU (95% CI) | 71,388.4 (62262.8–81851.6) | 61,296.9 (53962.1–69628.5) |
| Day 180 GMEU (95% CI) | 104,890.2 (93135.9–118128.0) | 94,026.0 (84606.1–104494.8) |
| GMFR from baseline to day 180 (95% CI) | 1.4 (1.3–1.6) | 1.5 (1.4–1.7) |
| SRR from baseline to day 180 (95% CI) | 6.0% (2.9–10.7) | 9.6% (5.7–14.9) |
| XBB.1.5 |  |  |
| Day 0 GMEU (95% CI) | 38,825.1 (33160.1–45457.9) | 32,881.9 (28434.7–38024.7) |
| Day 180 GMEU (95% CI) | 77,149.8 (68110.5–87388.8) | 61,647.7 (55459.2–68526.7) |
| GMFR from baseline to day 28 (95% CI) | 1.9 (1.7–2.2) | 1.9 (1.7–2.2) |
| SRR from baseline to day 180 (95% CI) | 21.4% (15.5–28.4) | 19.2 (13.7–25.8) |

ELISA=enzyme-linked immunosorbent assay; GMFR=geometric mean fold rise; GMEU=geometric mean ELISA units; GMEUR=ratio of GMEU between groups; IgG=immunoglobulin G; LLOQ=lower limit of quantification; SRR=seroresponse rate.

*An analysis of covariance with vaccine group as fixed effect and baseline value as covariate is performed to estimate the adjusted GMEU and GMEUR. The mean difference between vaccine groups and the corresponding CI limits is then exponentiated to obtain the ratio of GMEUs and the corresponding 95% CIs. Values <LLOQ are replaced by 0·5 $\times$ LLOQ for calculation of geometric means, GMFR, and GMEUR.

^†^Seroresponse is defined as a ≥4-fold increase from baseline values ≥LLOQ or ≥4-fold the LLOQ if the baseline value is <LLOQ.

**Table S6.** Anti-rS IgG responses against ancestral and XBB.1.5 SARS-CoV-2 strains by number of prior COVID-19 vaccinations (per-protocol analysis sets)

| Parameter | NVX-CoV2601 | | | | Bivalent | | |
| --- | --- | --- | --- | --- | --- | --- | --- |
| Prior COVID-19 vaccines | **2 Doses**  **(n=77)** | **≥3 Doses**  **(n=101)** | | **2 Doses**  **(n=89)** | | | **≥3 Doses**  **(n=105)** |
| Ancestral |  | | |  | | | |
| Day 0 GMEU (95% CI) | 47,256.5 (38795.0–57563.5) | 98,126.6 (82804.1–116284.3) | | 44,197.6 (36651.4–53297.4) | | | 81,234.8 (69167.2–95407.9) |
| Day 28 GMEU (95% CI) | 142,077.7 (119898.8–168359.3) | 219,256.7 (189362.0–253870.9) | | 143,182.9 (122188.9–167784.1) | | | 169,906.1 (146795.8–196654.6) |
| Day 28 adjusted GMEU* (95% CI) | 139,805.3 (121052.7–161462.9) | 207,278.3 (185453.5–231671.5) | | 145,194.3 (126991.0–166006.9) | | | 179,340.5 (160803.2–200014.8) |
| GMEUR* between groups (95% CI) | – | – | | 1.0 (0.85–1.26) | | | 0.9 (0.74–1.01) |
| GMFR from baseline to day 28 (95% CI) | 3.0 (2.5–3.5) | 2.2 (1.9–2.6) | | 3.2 (2.7–3.9) | | | 2.1 (1.9–2.4) |
| SRR^†^ from baseline to day 28 (95% CI) | 39.0 (28.0–50.8) | 18.8 (11.7–27.8) | | 33.7 (24.0–44.5) | | | 14.3 (8.2–22.5) |
| SRR difference between groups (95% CI) | – | – | | −5.3 (−19.8 to 9.4) | | | −4.5 (−15.0 to 5.7) |
| XBB.1.5 |  | |  | | |  | |
| Day 0 GMEU (95% CI) | 24,893.0 (20279.9–30555.5) | 54,902.4 (44379.4–67920.6) | | 24,699.8 (19803.2–30807.2) | | | 41,848.6 (34718.2–50443.5) |
| Day 28 GMEU (95% CI) | 126,626.7 (104407.0–153575.1) | 171,144.8 (146253.9–200271.7) | | 112,373.2 (94378.1–133799.4) | | | 113,593.2 (98467.4–131042.6) |
| Day 28 adjusted GMEU* (95% CI) | 126,435.0 (106533.0–150054.9) | 161,032.5 (142423.3–182073.1) | | 112,520.6 (95950.0–131952.9) | | | 120,446.7 (106781.8–135860.3) |
| GMEUR* between groups (95% CI) | – | – | | 0.9 (0.70–1.12) | | | 0.7 (0.63–0.89) |
| GMFR from baseline to day 28 (95% CI) | 5.1 (4.1–6.3) | 3.1 (2.6–3.7) | | 4.5 (3.7–5.7) | | | 2.7 (2.3–3.1) |
| SRR^†^ from baseline to day 28 (95% CI) | 61.0 (49.2–72.0) | 35.6 (26.4–45.8) | | 49.4 (38.7–60.2) | | | 28.6 (20.2–38.2) |
| SRR difference between groups (95% CI) | – | – | | −11.6 (−26.2 to 3.6) | | | −7.1 (−19.7 to 5.7) |

ELISA=enzyme-linked immunosorbent assay; GMFR=geometric mean fold rise; GMEU=geometric mean ELISA units; GMEUR=ratio of GMEU between groups; IgG=immunoglobulin G; LLOQ=lower limit of quantification; SRR=seroresponse rate.

*An analysis of covariance with vaccine group as fixed effect and baseline value as covariate is performed to estimate the adjusted GMEU and GMEUR. The mean difference between vaccine groups and the corresponding CI limits is then exponentiated to obtain the ratio of GMEUs and the corresponding 95% CIs. Values <LLOQ are replaced by 0·5 $\times$ LLOQ for calculation of geometric means, GMFR, and GMEUR.

^†^Seroresponse is defined as a ≥4-fold increase from baseline values ≥LLOQ or ≥4-fold the LLOQ if the baseline value is <LLOQ.

**Table S7.** Pseudovirus neutralising antibody response comparison of NVX-CoV2601 and NVX-CoV2373 (per-protocol analysis sets)

| **Parameter** | **NVX-CoV2601**  **(n=178)** | **NVX-CoV2373***  **(n=114)** |
| --- | --- | --- |
| **Ancestral** |  |  |
| Day 0 GMT (95% CI) | 1322.20 (1122.43–1557.54) | 202.47 (147.62–227.70) |
| Day 28 GMT (95% CI) | 3510.84 (3099.99–3976.14) | 4200.21 (3482.55–5065.75) |
| Day 28 adjusted GMT^†^ (95% CI) | 3000.29 (2613.30–3444.60) | 5368.24 (4485.53–6424.67) |
| GMTR^†^ between groups (95% CI) | 0.6 (0.44–0.71) | – |
| GMFR from baseline to day 28 (95% CI) | 2.7 (2.3–3.1) | 20.7 (14.8–29.0) |
| SRR^‡^ from baseline to day 28 (95% CI) | 34.8% (27.9–42.3) | 80.7% (72.3–87.5) |
| SRR difference between groups (95% CI) | −45.9% (−55.3 to −35.1) | – |
| **XBB.1.5** |  |  |
| Day 0 GMT (95% CI) | 208.40 (166.20–261.32) | 20.49 (19.12–21.97) |
| Day 28 GMT (95% CI) | 2533.08 (2107.17–3045.07) | 114.41 (95.34–137.30) |
| Day 28 adjusted GMT^†^ (95% CI) | 2004.32 (1658.99–2421.54) | 164.91 (128.10–212.28) |
| GMTR^†^ between groups (95% CI) | 12.2 (8.50–17.38) | – |
| GMFR from baseline to day 28 (95% CI) | 12.2 (9.5–15.5) | 5.6 (4.6–6.7) |
| SRR^‡^ from baseline to day 28 (95% CI) | 70.8% (63.5–77.3) | 38.6% (29.6–48.2) |
| SRR difference between groups (95% CI) | 32.2% (20.7–42.9) | – |

GMFR=geometric mean fold rise; GMT=geometric mean titre; GMTR=geometric mean titre ratio; LLOQ=lower limit of quantification; SRR=seroresponse rate.

*The 2019nCoV-301 phase 3 study included a pediatric expansion group of adolescents aged 12 to <18 years who received two primary series doses of NVX-CoV2373 and a 3^rd^ dose within 5 months of the primary series.

^†^An analysis of covariance with vaccine group as fixed effect and baseline value as covariate is performed to estimate the adjusted GMT and GMTR. The mean difference between vaccine groups and the corresponding CI limits is then exponentiated to obtain the ratio of GMTs and the corresponding 95% CIs. Values <LLOQ are replaced by 0·5 $\times$ LLOQ for calculation of geometric means, GMFR, and GMTR.

^‡^Seroresponse is defined as a ≥4-fold increase from baseline values ≥LLOQ or ≥4-fold the LLOQ if the baseline value is <LLOQ.

**2019nCoV-314 Study Investigators**

| **First names** | **Surnames** | **Institution** |
| --- | --- | --- |
| Carlos | Fierro | Johnson County Clinical Trials |
| Suchet | Patel | Velocity Clinical Research |
| Stephan | Sharp | Clinical Research Associates, Inc |
| Lawrence | Chu | Benchmark Research |
| Terry | Poling | AMR |
| Jeffrey | Adelglass | Research Your Health |
| Matthew | Braddock | Westside Center for Clinical Research |
| Kenneth | Etokhana | Tekton Research |
| Khozema | Palanpurwala | DM Clinical Research |
| Robyn | Hartvickson | Alliance for Multispecialty Research, LLC (AMR) |
| Wieslaw | Jakubowski | South Texas Clinical Research |
| Jibran | Atwi | Velocity Clinical Research |
| Marian | Shaw | Velocity Clinical Research |
| Doug | Logan | Velocity Clinical Research |
| Jacqueline | Alvarez | ITB Research |
| Darek | Eggleston | Mountain View CCT Research |
| Ausberto B | Hidalgo | Alfa Medical Research |
| Divina | Roman | Tekton Research |
| Erika | Clayton | DM Clinical Research - Chicago |
| John | Adams | Medical Colleagues of Texas, LLP |
